# Incidence of Arterial and Venous Thromboembolism in Cancer Patients

**DOI:** 10.1101/2023.05.17.23290143

**Authors:** Hai-Wei Deng, Jie Li, Wei-Yi Mei, Xiao-Xiong Lin, Qing Xu, Yuan-Sheng Zhai, Qian Zheng, Jin-Sheng Chen, Zhi-Bin Huang, Yun-Jiu Cheng

## Abstract

**Background and Aims:** The reported incidence of arterial thromboembolism (ATE) and venous thromboembolism (VTE) after cancer varies. A meta-analysis was performed to define the incidence of thromboembolism (TE) in cancer patients.

**Methods:** Articles were searched in PubMed and Embase from inception to November 1, 2022. Studies reporting the incidence data or data from which incidence could be estimated among patients with cancer and the explicit follow-up duration were included.

**Results:** Seventy-four studies involving 5059134 cancer patients were identified. The incidence rate per 1000 person-years was 11.60 (95% CI 7.62-15.58) for ATE, 6.11 (95% CI 3.70-8.53) for myocardial infarction, 9.07 (95% CI 7.48-10.66) for ischemic stroke, 2.11 (95% CI 0.89-3.31) for another ATE, 26.32 (95% CI 24.46-28.18) for VTE, 12.69 (95% CI 11.51-13.87) for deep vein thrombosis, 5.94 (95% CI 5.29-6.59) for pulmonary embolism, and 13.18 (95% CI 9.93-16.42) for another VTE. In addition, the highest incidence of ATE was observed in patients with gastrointestinal cancer, while patients with pancreatic cancer had the highest incidence of VTE. The risk of ATE and VTE increased at the initial stage of cancer, and then declined and became non-significant.

**Conclusion:** This meta-analysis provided overall estimates of ATE and VTE incidence in cancer patients, adding an important insight into the trajectory of the development of TE in cancer patients, which could help reduce the risk of TE in cancer patients in the future.

## Background

Cancer is a leading cause of mortality worldwide. The recent decrease in mortality from various types of cancers has been attributed to advances in cancer screening and diagnosis, as well as improvement in cancer treatments [1, 2]. However, improved survival leads to cardiovascular complications that are common after a cancer diagnosis. Patients with cancer are prone to develop arterial thromboembolism (ATE) and venous thromboembolism (VTE). Importantly, thromboembolism (TE) in cancer patients posed a serious threat to public health. TE increases mortality, disrupts cancer treatments, and causes a significant economic burden [3-5].

There are several reports on the incidence of VTE in patients with different types of cancer. An American retrospective cohort study in the USA showed that VTE occurred in 13.9% of lung cancer patients.[6] Chew and colleagues reported a 2-year cumulative VTE incidence of 1.2% among 108255 patients with breast cancer [7]. Additionally, Ziegler found that 15 of 719 acute leukemia patients (2.09%) had VTE [8]. The incidence of ATE in cancer patients has also been documented. A prospective observational cohort study involving 1880 cancer patients found that 2.6% of patients developed ATE [9]. Despite much work already conducted in this field, no large-scale systematic studies have been performed to investigate the incidence of TE in cancer patients, especially ATE.

Given this background, an up-to-date understanding of the incidence of ATE and VTE in cancer patients is needed. This would help clinicians attach more importance to TE awareness among cancer patients. Therefore, a meta-analysis of published studies was conducted to ascertain the incidence of ATE and VTE in cancer patients and perform a further analysis based on the type of cancer. The relationship between the duration of follow-up and incidence of TE after cancer diagnosis was also explored.

## Materials and Methods

This meta-analysis was conducted in accordance with the Meta-analyses and Systematic reviews of Observational Studies (MOOSE). The protocol was registered in the Prospective Register of Systematic Reviews (PROSPERO CRD42021272276) before data extraction.

### Search strategy and selection criteria

Articles were searched in PubMed and Embase from inception to November 1, 2022, using a combination of the following text and key words, both as Medical Subject Heading (MeSH) terms and text words: “cancer”, “carcinoma”, “artery”, “vein”, “thromboembolism”, and “thrombosis”. The search was limited to humans. No restrictions were applied based on language, sex, location, or duration of follow-up. The reference list of all relevant articles and reviews was manually searched to identify other relevant articles. In the case of multiple reports on the same population or subpopulation, only studies reporting the most recent or informative data were considered.

Studies were considered eligible if they reported the incidence data or data from which incidence could be estimated (numerator and denominator) among cancer patients and explicit follow-up duration. Articles were excluded if they were cross-sectional studies, review articles, commentaries, case reports or editorials. Studies involving a population receiving a specific or single treatment and primary studies with a small sample size (<100 participants) were also excluded.

Two investigators (Hai-Wei Deng and Jie Li) independently screened the titles and abstracts of retrieved articles. Full-text articles were screened for inclusion by the same reviewers. Selection discrepancies were resolved through discussion and consensus.

### Data analysis

Relevant data were extracted by two investigators (Hai-Wei Deng and Jie Li) using a preconceived and standardised data extraction form. The following data were abstracted from included studies: first author, year of publication, region/country, data sources, study type, study design, baseline patient characteristics, sample size, follow-up data, and outcomes. The primary outcome of this analysis was the incidence of TE events, including ATE and VTE. ATE was defined as myocardial infarction (MI), ischemic stroke (IS), and peripheral artery disease; VTE was defined as deep vein thrombosis (DVT), pulmonary embolism (PE), and unusual site thrombosis (i.e. splanchnic vein or cerebral vein). Additionally, the total ATE events included MI and IS at least, while the total VTE events had at least DVT and PE.

We did meta-analyses in Stata software, version 15.0 (Stata Corp, College Station, TX) to summarise reported data across studies. The incidence of ATE and VTE in cancer patients was expressed as per 1000 person-years of follow-up. Weighted meta-analytic incidence estimates for outcomes with the variance-stabilizing Freeman-Tukey double-arcsine transformation were calculated. We used an inverse-variance random-effects model because of a wide range of settings in different populations in studies. All estimates were reported with their 95% confidence intervals (CIs). Moreover, because CIs based on random-effects models are wider than those from fixed-effects models, random-effects results are conservative or “worst-case scenario”. Heterogeneity was assessed using the I^2^ (95% CI) statistic (0-25%: minimal heterogeneity; 26-75%: moderate heterogeneity; >75%: substantial heterogeneity) [10].

Subgroup analysis based on study design, age, region, and sample size was performed. Meta-regression was used to calculate p-values between subgroups. Further, a fractional polynomial model was used to examine the relationship between follow-up years and the incidence of TE after cancer diagnosis.

A funnel chart and Egger’s statistical tests were used to evaluate publication bias [11]. All statistical tests conducted were two-sided, and values of p < 0.05 were considered statistically significant.

## Results

A total of 58334 records were initially identified through a comprehensive search, of which 10035 articles were retained after the elimination of duplicates and screening of the titles. Of these, 232 reports underwent full-text screening after screening of the abstracts. A total of 74 studies were finally included in the meta-analysis (Supplement Figure S1).

A total of 5059134 individuals were examined (ranging from 195 to 2689670 in each study and approximately 3.56 million from North America, 0.76 million from Asia, and0.74 million from Europe). Multiple tumor types, including lung, esophagus, gastrointestinal tract, colorectum, liver, pancreas, kidney, bladder, prostate, testis, breast, uterus, ovary, brain, head and neck, and thyroid cancers, malignant melanoma, lymphoma, leukaemia, multiple myeloma, etc., were examined in various studies. The duration of follow-up across studies ranged from 0.17 to 30 years. Fourteen of the studies were prospective and sixty were retrospective. Studies were published between 2005 and 2021. The characteristics of the included studies are summarised in supplementary, Supplement Table S1.

### Incidence of ATE

ATE in cancer patients was reported in 19 of 74 studies (Figure 1), with data for 3335008 individuals. The incidence rate varied around 72 times, from 0.76 (95% CI 0.67-0.87) to 54.92 (95% CI 31.69-93.53) per 1000 person-years. The overall pooled incidence of ATE in cancer patients was 11.60 per 1000 person-years (95% CI 7.62-15.58) with very high between-sample heterogeneity (p < 0.01; I^2^ = 99.98%). The funnel plot is shown in Supplement Figure S12. Egger’s test did not indicate publication bias (p = 0.149).

**Figure 1.**
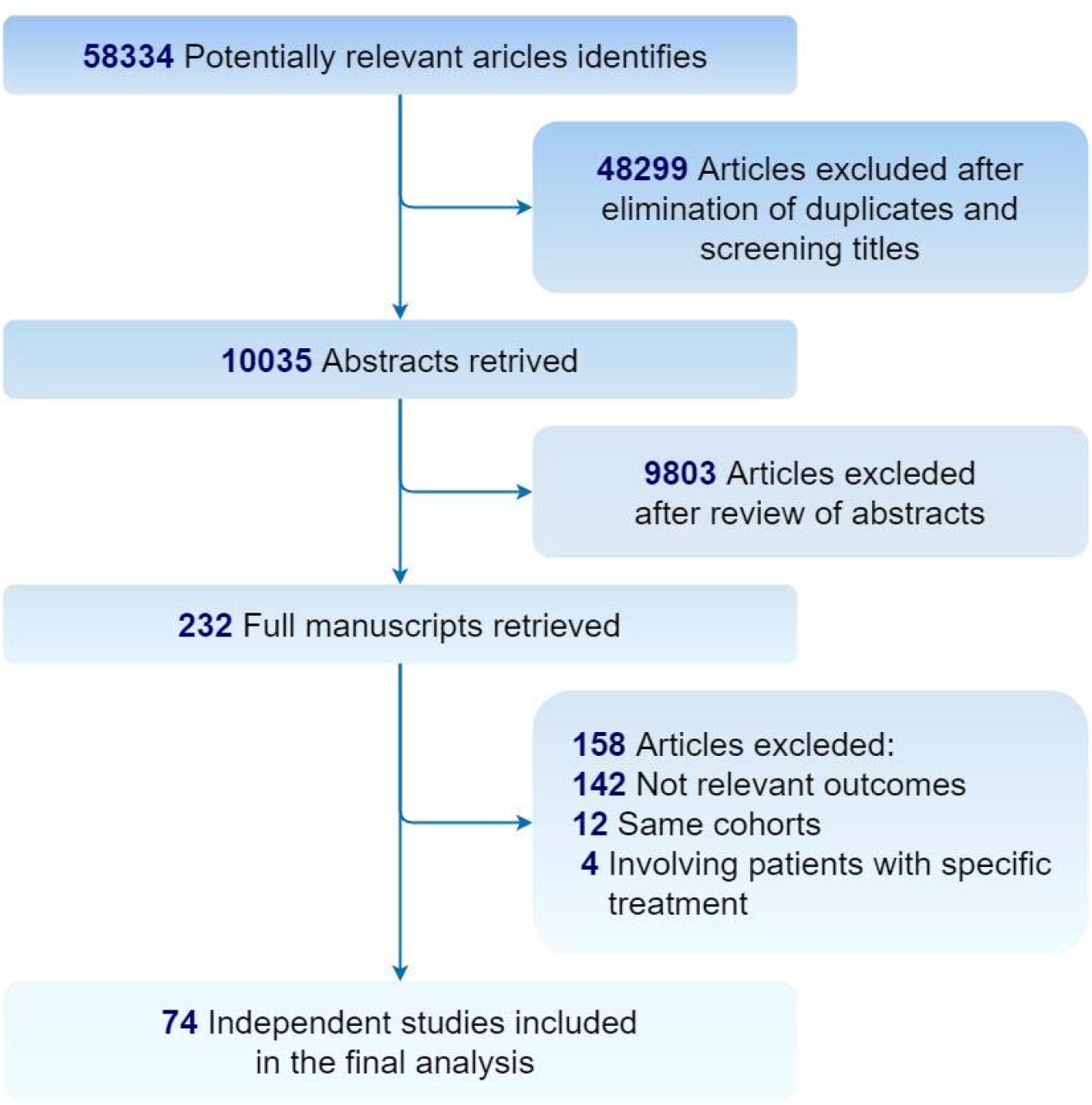
Flowchart of the selection of studies included in meta-analysis.

The incidence rate of MI was reported in 15 studies involving 3254612 patients. Incidence varied almost 46 times, from 0.35 (95% CI 0.30-0.41) to 16.03 (95% CI 13.08-19.64) per 1000 person-years. The overall incidence was 6.11 per 1000 person-years (95% CI 3.70-8.53; Supplement Figure S2), with high interstudy heterogeneity (p < 0.01; I^2^ = 99.96%). The incidence rate of IS was reported in 26 studies involving 3707421 participants and varied from 0.50 (95% CI 0.44-0.58) to 41.19 (95% CI 21.82-76.41) per 1000 person-years. Pooled incidence was 9.07 per 1000 person-years (95% CI 7.48-10.66; Supplement Figure S3), with high heterogeneity (p < 0.01; I^2^ = 99.96%). The overall incidence of another ATE was 2.11 per 1000 person-years (95% CI 0.89-3.31; Supplement Figure S4).

### Incidence of VTE

The pooled incidence of VTE in cancer patients from 52 studies with 1563743 subjects was 26.32 per 1000 person-years (95% CI 24.46-28.18; Figure 2) using a random-effects model because heterogeneity was present (I^2^ = 99.85%, p < 0.01). The incidence estimates ranged from 0.62 (95% CI 0.55-0.70) to 223.55 (95% CI 193.65-256.59) per 1000 person-years. Graphical visualisation of the funnel plot (Supplement Figure S13) and Egger’s test (p<0.01) suggested the presence of publication bias.

**Figure 2.**
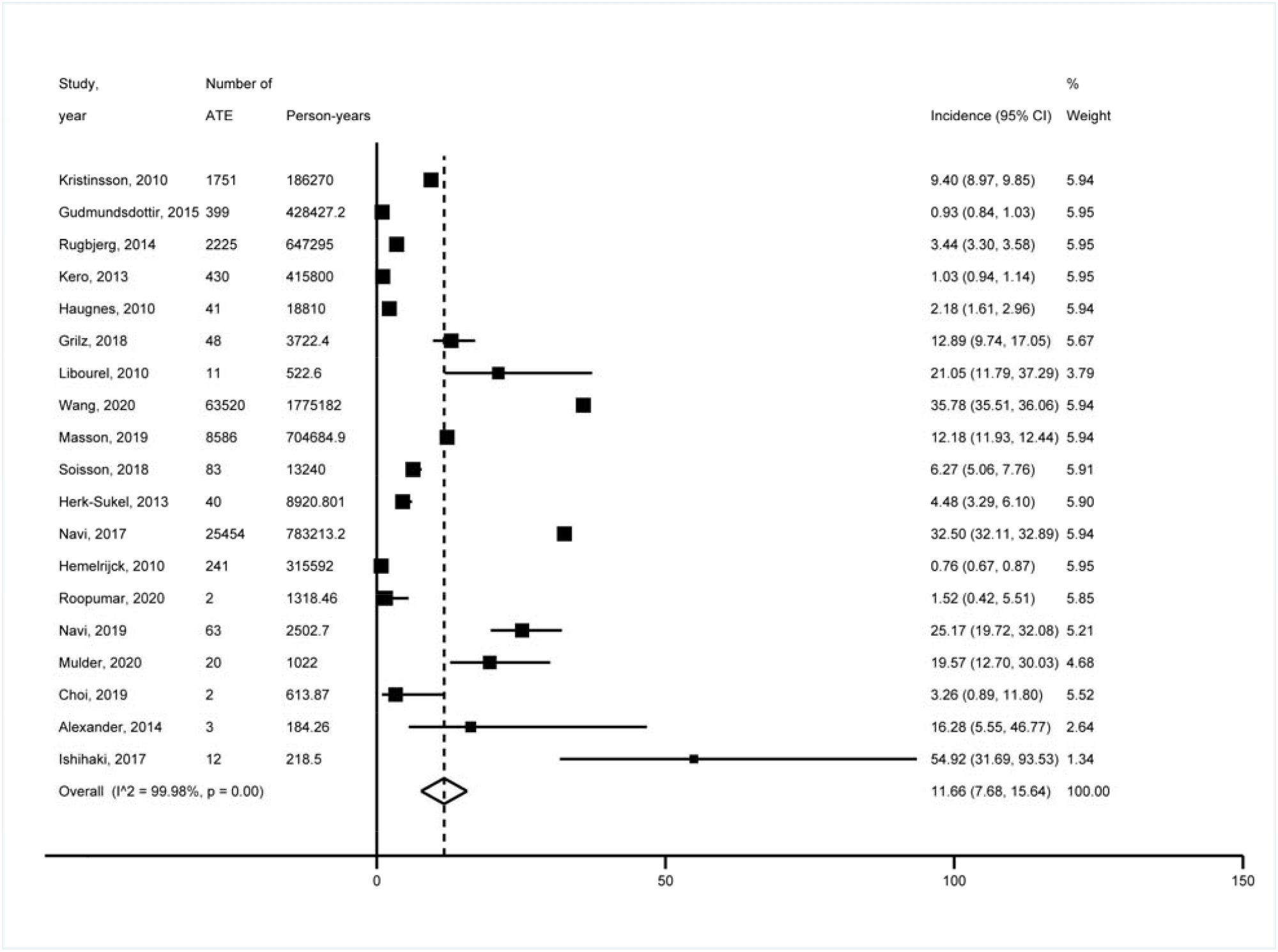
Forest plot showing the incidence of ATE.

DVT was reported in 29 studies involving 934782 patients. The incidence rate ranged from 1.69 (95% CI 1.55-1.84) to 102.94 (95% CI 95.91-110.43) per 1000 person-years. The overall incidence of DVT in cancer patients was 12.69 per 1000 person-years (95% CI 11.51-13.87; Supplement Figure S5), with very high heterogeneity (p < 0.01; I^2^ = 99.96%). Meta-analysis of 31 studies describing 966828 cancer patients revealed that the incidence of PE was 5.94 per 1000 person-years (95% CI 5.29-6.59; Supplement Figure S6), with high heterogeneity (p < 0.01; I^2^ = 99.30%). The incidence rate varied around 263 times, from 0.26 (95% CI 0.21-0.31) to 68.49 (95% CI 48.40-96.09) per 1000 person-years. Additionally, the pooled incidence of another VTE was 13.18 per 1000 person-years (95% CI 9.93-16.42; Supplement Figure S7).

### Stratified analysis

To further explore the relationship between cancer types and the incidence of ATE and VTE, we performed an analysis stratified by type of cancer. The results of the correlational analysis are shown in Table1 and Table2. Patients with gastrointestinal cancer (55.08 per 1000 person-years (95% CI 35.45-75.70)), pancreatic cancer (46.89 (95% CI 14.43, 79.36)), and bladder cancer (37.03 (95% CI 35.37, 38.70)) ranked as the top three in terms of the incidence of ATE. The highest pooled incidence of MI was observed in patients with gastrointestinal cancer (30.77 per 1000 person-years (95% CI 27.92-33.61)). The incidence of IS was highest in lung cancer patients (33.66 per 1000 person-years (95% CI 10.72-56.60)).

**Table 1.** Analysis of ATE stratifying by type of cancer

| Type of cancer | ATE | MI | IS |
| --- | --- | --- | --- |
|  | per 1000 person-years<br>(95% confidence<br>interval) | per 1000 person-years<br>(95% confidence<br>interval) | per 1000 person-years<br>(95% confidence<br>interval) |
| Bladder | 37.03 (35.37, 38.70) | 14.86 (13.83, 15.97) | 2.35 (2.04, 2.66) |
| Breast | 19.54 (5.32, 33.75) | 9.22 (4.73, 13.71) | 9.95 (5.30, 14.60) |
| Brain | 36.22 (0, 104.53) | 1.15 (0.91, 1.39) | 22.63 (18.32, 26.94) |
| Colorectum | 29.39 (2.90, 55.87) | 12.85 (1.37, 24.33) | 19.36 (2.49, 36.22) |
| Esophagus | 19.57 (12.70, 30.03) | 2.94 (1.00, 8.59) | 5.87 (2.69, 12.75) |
| Gastrointestinal tract | 55.08 (35.45, 74.70) | 30.77 (27.92, 33.61) | 15.93 (11.28, 20.58) |
| Head and neck | 38.31 (10.57, 129.35) | / | 4.05 (3.65, 4.45) |
| Kidney | 28.11 (3.02, 53.19) | 12.81 (0, 28.31) | 9.22 (0, 20.51) |
| Leukaemia | 7.70 (0, 17.55) | 0.31 (0.14, 0.47) | 1.00 (0.22, 1.79) |
| Liver | 19.13 (17.38, 20.87) | 11.45 (10.09, 12.81) | 5.75 (4.78, 6.71) |
| Lung | 35.72 (6.45, 64.99) | 21.33 (5.98, 36.67) | 33.66 (10.72, 56.60) |
| Lymphoma | 24.55 (3.99, 45.11) | 12.80 (2.81, 22.79) | 12.23 (8.54, 15.91) |
| Malignant melanoma | 0.73 (0.50, 1.08) | 0.41 (0.24, 0.69) | 0.23 (0.17, 0.29) |
| Multiple myeloma | 12.19 (4.89, 19.49) | 6.79 (6.41, 7.16) | 3.23 (2.98, 3.49) |
| Ovary | 21.55 (3.81, 112.45) | / | 1.69 (1.38, 2.00) |
| Pancreas | 46.89 (14.43, 79.36) | 22.40 (8.61, 36.19) | 27.34 (9.58, 45.11) |
| Prostate | 20.89 (5.31, 36.47) | 14.56 (9.44, 19.68) | 13.80 (8.54, 19.07) |
| Testis | 1.27 (0.95, 1.60) | 0.72 (0.48, 0.97) | 0.54 (0.46, 0.63) |
| Thyroid | 0.76 (0.54, 1.08) | 0.25 (0.13, 0.45) | 0.74 (0.58, 0.90) |
| Uterus | 31.35 (5.56, 157.88) | / | / |

ATE: Arterial Thromboembolism; MI: Myocardial Infarction; IS: Ischemic Stroke

**Table 2.** Analysis of VTE Stratifying by Type of Cancer

| Type of cancer | VTE | DVT | PE |
| --- | --- | --- | --- |
|  | per 1000 person-years<br>(95% confidence<br>interval) | per 1000 person-years<br>(95% confidence<br>interval) | per 1000 person-years<br>(95% confidence<br>interval) |
| Bladder | 19.65 (7.83, 31.47) | 7.57 (6.89, 8.26) | 1.68 (1.36, 2.01) |
| Breast | 11.32 (8.54, 14.11) | 3.50 (2.10, 4.91) | 2.25 (1.30, 3.20) |
| Brain | 45.40 (23.57, 67.23) | 12.21 (10.97, 13.45) | 2.20 (1.67, 2.73) |
| Colorectum | 24.24 (17.71, 30.77) | 5.48 (4.93, 6.04) | 4.46 (0, 9.84) |
| Esophagus | 25.13 (4.75, 45.50) | 2.95 (2.17, 3.73) | 0.36 (0.09, 0.63) |
| Gastrointestinal tract | 28.94 (20.64, 37.24) | 12.03 (6.23, 17.83) | 8.49 (2.78, 14.19) |
| Head and neck | 2.43 (2.12, 2.78) | 2.17 (1.88, 2.51) | 0.33 (0.23, 0.47) |
| Kidney | 16.60 (9.24, 23.96) | 5.05 (4.34, 5.77) | 1.54 (1.14, 1.93) |
| Leukaemia | 17.90 (12.41, 23.39) | 4.97 (3.25, 6.69) | 2.57 (1.28, 3.87) |
| Liver | 10.34 (5.83, 14.84) | 4.81 (4.45, 5.17) | 0.65 (0.52, 0.78) |
| Lung | 40.09 (34.31, 45.87) | 35.04 (27.49, 42.58) | 22.96 (16.44, 29.48) |
| Lymphoma | 32.83 (25.44, 40.21) | 16.11 (11.62, 20.60) | 9.36 (5.23, 13.49) |
| Malignant melanoma | 17.11 (15.86, 18.35) | / | / |
| Multiple myeloma | 19.85 (8.67, 31.03) | 9.55 (2.87, 16.24) | 2.90 (0.54, 5.62) |
| Ovary | 34.51 (24.44, 44.59) | 12.87 (4.77, 20.97) | 11.32 (0, 25.74) |
| Pancreas | 55.07 (39.28, 70.86) | 10.01 (3.99, 16.03) | 9.96 (1.18, 18.74) |
| Prostate | 10.92 (6.93, 14.90) | 4.36 (2.56, 6.16) | 2.66 (1.60, 3.72) |
| Testis | 6.06 (2.39, 9.74) | 4.75 (2.30, 9.77) | 0.68 (0.12, 3.83) |
| Thyroid | 12.29 (9.77, 15.44) | / | / |
| Uterus | 20.37 (9.40, 31.35) | 9.68 (8.73, 10.73) | 3.77 (3.19, 4.45) |

VTE: Venous Thromboembolism; DVT: Deep Vein Thrombosis; PE: Pulmonary Embolism

Inspection of the data revealed that pancreatic cancer patients had the highest incidence of VTE (55.07 per 1000 person-years (95% CI 39.28-70.06)), followed by those with brain cancer (45.40 (95% CI 23.57-67.23)) and lung cancer (40.09 (95% CI 34.31-45.87)). Patients with lung cancer had the highest incidence of DVT (35.04 per 1000 person-years (95% CI 27.49-42.58)) and PE (22.96 (95% CI 16.44 -29.48)).

To explore the possible causes of this heterogeneity, we performed stratified analyses across several key study characteristics and clinical factors, including study design (prospective or retrospective), region (Asia, Europe, or North America), age (<60, 60-69, or ≥70), and sample size (<10000, 10000-99999, or ≥100000). In the ATE subgroup, differences in study design, region, age, and sample size were not sources of heterogeneity (Supplement Figure S8). However, age and sample size influenced the incidence of VTE (Supplement Figure S9). Patients aged <60 years had the highest incidence of VTE after cancer (81.08 per 1000 person-years (95% CI 65.76, 96.41)), followed by those aged 60-69 years (61.36 (54.44, 68.28)) and 70 years or older (12.05 (8.90, 15.19)). Besides, a higher incidence of VTE was found in studies with small sample sizes.

### Follow-up duration and incidence of TE

Further analysis was conducted to determine the relationship between ATE incidence and cancer follow-up duration. The follow-up was <5 years in 25 studies, 5-10 years in 2 studies, and ≥10 years in 5 studies (Supplement Figure S10). The graph showed a steep increase in the incidence of ATE at the initial stages of cancer. Cancer was not associated with an increased risk for developing ATE after a longer duration of follow-up (approximately >10 years) (Figure 3 (A)). Similarly, several studies contained data on VTE incidence in relation to follow-up duration (Supplement Figure S11). The same trend was observed for VTE incidence, as illustrated in Figure 3 (B).

**Figure 3.**
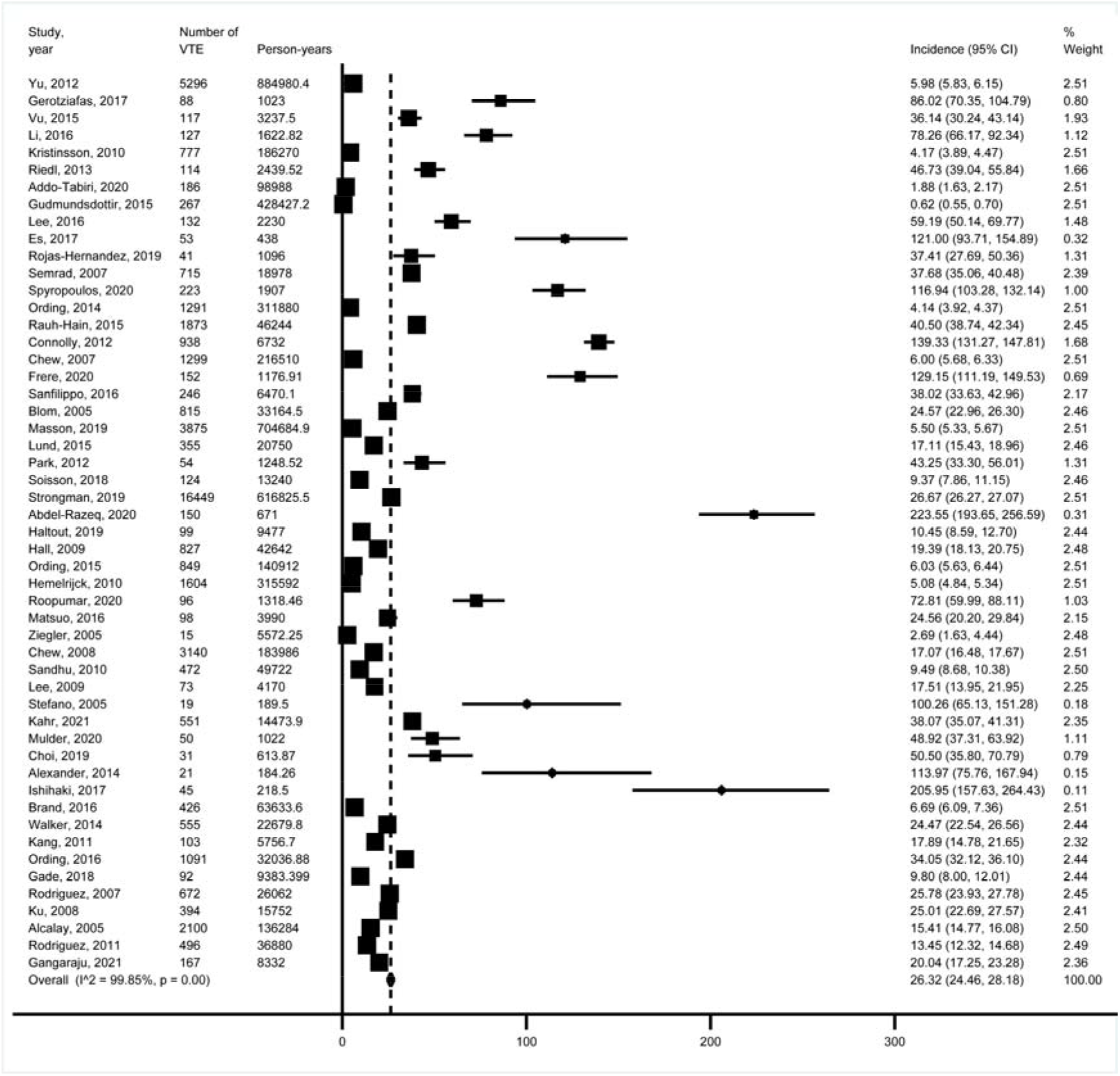
Forest plot showing the incidence of VTE.

**Figure 4.**
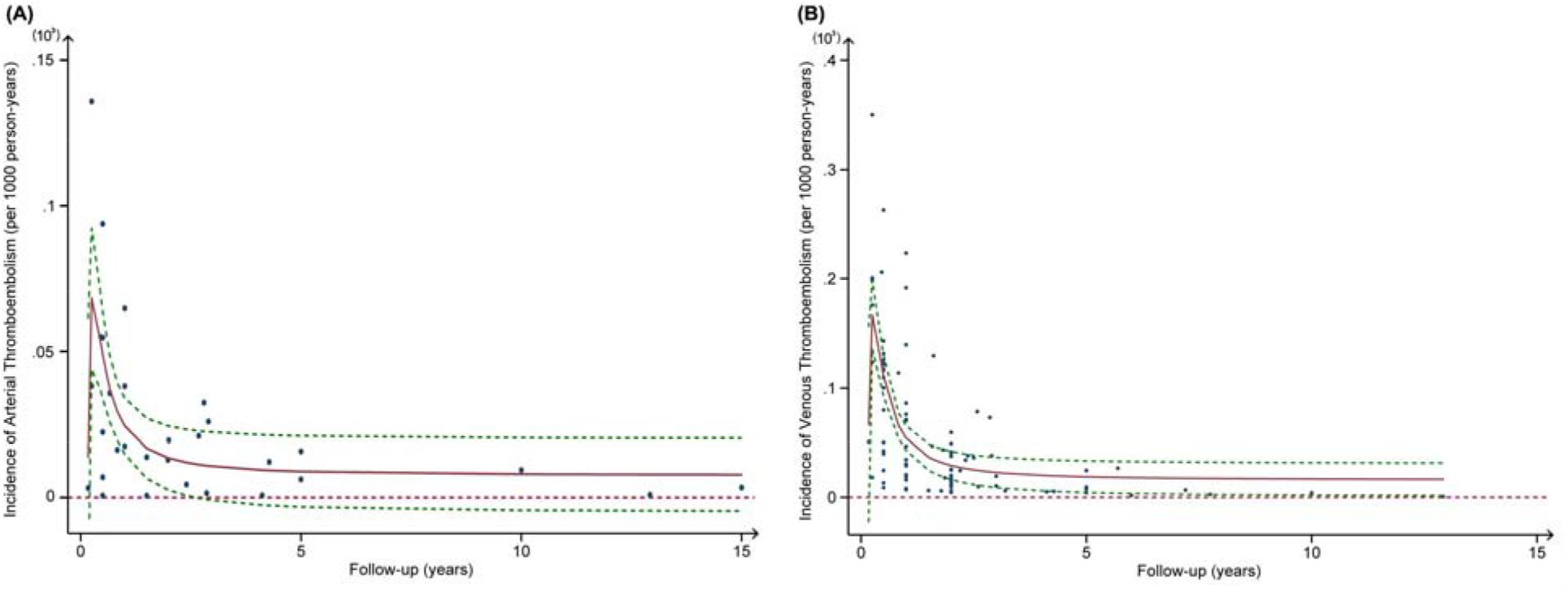
The relationship between thromboemlism incidence and cancer follow-up duration.

## Discussion

The present meta-analysis of 74 studies found that the incidence per 1000 person-years was 11.60 for ATE, 6.11 for MI, 9.07 for IS, 2.11 for another ATE, 26.32 for VTE, 12.69 for DVT, 5.94 for PE, and 13.18 for another VTE.

The relationship between malignancy and TE has been extensively studied for years. Nowadays, many cancer survivors remain at high risk of thromboembolic events, including both ATE and VTE [12]. The incidence of ATE and VTE varied extremely between studies. Kristinsson et al. reported that among 18627 multiple myeloma (MM) patients, 1751 had ATE (9.40 per 1000 person-years (95% CI 8.97-9.85)) and 777 had VTE (4.17 per 1000 person-years (95% CI 3.89-4.47)) at 10 years after MM diagnosis in Japan [13]. In a retrospective analysis of the USA, the 6-month incidence of ATE was 4.7% (32.50 per 1000 person-years (95% CI 32.11-32.89)) [14]. Analysis of a UK cohort showed that the incidence of VTE in all cancers was 13.9 per 1000 person-years (95% CI 13.4–14.4) [15]. Several factors could affect the results. Most previous studies focused solely on demographically homogenous populations. Thus, different sex, age, region, and ethnic groups were included in our study to provide a robust comprehensive analysis. In addition, a more detailed analysis of the incidence of MI, IS, DVT, and PE was presented in our study.

VTE is associated with the alteration of blood flow, endothelial damage, and increase of procoagulant activity, while ATE is thought to be related to the atherosclerotic plaque rupture, facilitated by enhanced thrombogenicity. There are several possible explanations for the increased risk of TE in cancer patients. First, cancer is associated with a hypercoagulable state involving the activation of diverse hemostatic components, such as platelets, vascular endothelium, coagulation and fibrinolytic pathways, and monocytes [16, 17]. Increased plasma viscosity and immobility in cancer patients may lead to stasis and the development of thrombosis [18]. Cancer can activate the innate immune system that can initiate thrombosis through neutrophil extracellular trap formation [19]. Cancer treatments, including surgery, chemotherapy, and radiotherapy also contribute to TE [20, 21].

The absolute risk of ATE and VTE in cancer patients varies widely depending on the site of cancer. Among diverse cancer types, we found that patients with gastrointestinal cancer, pancreatic cancer, bladder cancer, brain cancer, and lung cancer had a higher risk of developing ATE. Although one unexpected finding was that head and neck cancer patients had a high incidence of ATE, the small sample size precludes any definitive conclusions to be drawn. A higher incidence of MI and IS was also found among patients with lung cancer, gastrointestinal cancer, and pancreas cancer. The incidence of VTE was higher in patients with pancreatic cancer, brain cancer, lung cancer, ovarian cancer, and lymphoma. Moreover, patients with lung cancer, lymphoma, and ovarian cancer had a higher risk of DVT and PE. The mechanism of increased TE in specific cancer types is under debate. The difference in TE incidence between cancer types appeare to be driven by common risk factors that cancer and TE shares, such as smoking and age. This may also because these cancer types have intrinsic properties of procoagulant activity. It is well known that aggressive cancers are more likely to cause coagulopathy. In addition, specific anti-cancer treatments can lead to an increased risk of TE, such as cisplatin, vascular endothelial growth factor (VEGF)/vascular endothelial growth factor receptor (VEGFR) inhibitors, L-asparaginase, thalidomide, lenalidomide, and tamoxifen [22-24].

Notably, the risk of ATE and VTE in cancer patients varied during the disease. The risk of ATE and VTE tended to increase during the initial stage after diagnosis of cancer, and then declined and became non-significant, which corroborates the findings of a great deal of the previous work. Navi et al. found that the risk of ATE was highest soon after cancer diagnosis,^14^ and Vu et al. found that the risk of VTE was greatest within the first year following diagnosis and declined with time after diagnosis in leukemia patients [25]. Brand et al. also found that the incidence of VTE was highest in the first 6 to 12 months after diagnosis in breast cancer patients [26]. This result may be explained by the fact that cancer activity and treatments are most intense in the early stage, and are then attenuated over time. Alternatively, it is possible that invasive surgeries and thrombocytopenia sometimes contribute to interruptions in anticoagulation or antiplatelet, which is prone to TE. Also, during the first months after cancer diagnosis patients are in close contact with many physicians, they probably received more detection in the early stage, which would lead to the rapid discovery of TE. Furthermore, it is more likely that physicians in the later course (especially in palliative patients) will attribute TE symptoms to the underlying cancer and not initiate further diagnostic testing.

This meta-analysis has some important implications. The results may motivate patients to attend screening examinations, especially patients with specific cancer types and at the initial stage. Clinicians should consider with care an optimal antithrombotic treatment strategy for primary prevention of TE in cancer patients. Therefore, it can help guide management and monitoring to reduce the future risk of TE in cancer patients.

This study has several strengths, including the strict inclusion criteria, using a relatively large size, the diversity of the study population, the multiple outcomes, and the relationship between follow-up duration and risk of TE.

However, this study also has some limitations. First, the study has the limitations of being a retrospective analysis. Second, considerable heterogeneity was observed in the results of various studies. Population characteristics, including age range, smoking habit, treatment strategy, presence or absence of hypertension, hyperglycaemia, dyslipidaemia, etc., might have led to the heterogeneity of the studies. Additionally, the study relies on published data and is subject to publication bias. Finally, limited data on individual participants make it difficult to identify the independent associations between individual variables and study outcomes. Further research is warranted to better understand the underlying mechanisms, evaluate the impact of potential risk factors, and determine the optimal antithrombotic treatment strategy for cancer patients.

## Conclusions

In summary, this meta-analysis provides overall estimates of ATE and VTE incidence rates in cancer patients (including the incidence of MI, IS, DVT, and PE). The study also showed that patients with gastrointestinal cancer, pancreatic cancer, bladder cancer, brain cancer, and lung cancer had a higher risk of developing ATE, while patients with pancreatic cancer, brain cancer, lung cancer, ovarian cancer, and lymphoma had a higher incidence of VTE. Furthermore, it was found that the risk of TE increased during the initial stage after diagnosis of cancer and then declined and became non-significant. This would help clinicians attach more importance to TE awareness, reducing the future risk of TE in cancer patients.

## Data Availability

The data that support the findings of this study are available from the corresponding author, upon reasonable request.

## ABBREVIATIONS AND ACRONYMS

ATE: Arterial Thromboembolism
MI: Myocardial Infarction
IS: Ischemic Stroke
VTE: Venous Thromboembolism
DVT: Deep Vein Thrombosis
PE: Pulmonary Embolism
CI: confidence interval

## Acknowledgements

None.

---

Fig1 Forest plot showing the incidence of ATE.

Fig2 Forest plot showing the incidence of VTE.

Fig3 The relationship between incidence and cancer follow-up.

FigS1 Flowchart of the selection of studies.

FigS2 Forest plot showing the incidence of MI.

FigS3 Forest plot showing the incidence of IS.

FigS4 Forest plot showing the incidence of another ATE.

FigS5 Forest plot showing the incidence of DVT.

FigS6 Forest plot showing the incidence of PE.

FigS7 Forest plot showing the incidence of another VTE.

FigS8 Subgroup analysis of ATE.

FigS9 Subgroup analysis of VTE.

FigS10 Forest plot showing the incidence of ATE with different follow-up.

FigS11 Forest plot showing the incidence of VTE with different follow-up.

FigS12 ATE funnel graph.

FigS13 VTE funnel graph.

